# Water, Health, and Social Technologies: One Million Cisterns Programme case study

**DOI:** 10.1101/2025.04.23.25326299

**Authors:** José Firmino de Sousa Filho, Walisson Araújo, Maraina Sebastião, Adalton Fonseca, Raíza Tourinho, Denise Pimenta, Gervásio Santos, Lucas Emanuel, Roberto F. S. Andrade, Gustavo Casais, Gisele Paixão, Andrea Ferreira, Rachel Coelho, Maria Yuri Ichihara, Júlia Pescarini, Joanna Guimarães, Paulo V. da Costa, Ismael Silveira, Rafael Silva, Rita Ribeiro, Maurício L. Barreto, the SEDHI team

## Abstract

**Background:** This paper focuses on the impacts of climate change on vulnerable ecosystems and its implications for the health and well-being of populations. It specifically examines the Semi-arid region of Brazil, where the introduction of a social climate adaptation tool, cisterns, has brought about significant positive changes. Cisterns, a low-cost climate adaptation technology, can be replicated globally, reducing the negative health impacts of frequent droughts, especially for vulnerable groups in remote rural areas.

**Objective:** We analyze the impact of the “One Million Cisterns Program” (P1MC) on health by synthesizing the literature and modeling its interactions with climatic and environmental factors with the DPSEEA framework.

**Methods:** Our case study employs a multidisciplinary approach, focusing on two key objectives: (i) synthesizing the literature on the implementation of the P1MC and its association with health outcomes; and (ii) developing a conceptual framework to model the relationship between climatic and environmental factors, adaptive ecosystems, and health outcomes. The *Driving Force–Pressure–State–Exposure–Effect–Action* (DPSEEA) framework evaluates the structural connections between climate change and human health.

**Findings:** The study found a significant gap in the literature concerning the relationship between P1MC and health outcomes. Cisterns target the pressure/state linkages related to contextual factors and health effects, addressing the root causes of drought-related health issues. This framework also provides a foundation for collaboration among health, environmental, and policy sectors to address shared challenges, such as water security and health outcomes.

**Conclusion:** We offer a multidisciplinary analytical framework that can be used to explore various perspectives – environmental, social, and health-related – with experts and stakeholders to develop and improve adaptive social technology strategies for living in the emergence of climate change. This framework also facilitates the implementation of qualitative and quantitative well-being and health assessments.

## 1. Background

The Brazilian Semi-arid expanse ranks among the largest and most densely populated in the world[1]. In 2021, the Federal Government released the latest demarcation, revealing a notable augmentation in the domain’s municipal coverage and territorial stretch[2]. Currently, the Semi-arid domain constitutes 15.5% of the national territory, encompassing approximately 1,427 municipalities and accommodating 30.3 million people (equivalent to 14.6% of the total Brazilian population and 54.7% of the Northeastern region’s inhabitants).

In addition to water scarcity and prolonged droughts exacerbated by climate change, which hinders access to water for consumption and food production, the Semi-arid region harbors the largest proportion of the rural Brazilian population facing poverty and extreme poverty. Approximately 47% of the region’s population experienced severe food insecurity immediately following the COVID-19 pandemic, with socioeconomic, gender, and racial inequities becoming further pronounced[3].

The population of the Semi-arid region is racially, ethnically, and culturally diverse, featuring the Black population, indigenous, and *quilombola* communities in the area. These social groups understand nature from different perspectives than those so-called Westerns. Traditional communities comprehend land, water, and animals through their cosmologies, avoiding accepting the earth only as an economic resource[4]. Most of this population is engaged in subsistence farming and livestock activities[5]. Nevertheless, despite their cultural and identity richness, they are particularly exposed to stigmas, harmful environmental stressors, and socially deprived areas, which increase their vulnerability to extreme weather events due to climate change[6].

In the Semi-arid region, resource conservation is crucial. It involves storing water for drinking, agriculture, and livestock, as well as preserving food and seeds. The NGO *Articulação do Semiárido* (*ASA*) was formed to improve coexistence in the Brazilian Semi-arid region. The ASA is a network of civil society organizations that works for the social, economic, political, and cultural development of the Semi-arid region of Brazil, bringing together hundreds of entities, including other NGOs, rural workers’ unions, farmers’ associations, cooperatives, environmental organizations, pastoral groups, churches, and others[7].

As communities engage with the ASA association and in constructing cisterns, they learn the power of collective action in asserting their rights, fostering community engagement, and promoting full citizenship. We present a detailed cultural and territorial perspective in the supplementary material (SM1). The P1MC experience informs public policy, emphasizing grassroots involvement and integrating local insights into policy formulation. By mobilizing society, the program advocates for effective and inclusive policies that uphold the rights of rural populations to access clean water[8].

Over the past two decades, access to potable water in the Semi-arid region has evolved from emergency relief to a government-endorsed policy with national budget allocations. The National Council for Food and Nutritional Security (Consea) has rightly recognized P1MC cisterns as a lifeline for water and food security in the region[8]. The provision of potable water is essential for food security and other human rights. The region’s water scarcity is a result of limited rainfall and social factors like unequal distribution and ownership[9,10]. Traditional drought mitigation measures, such as large reservoirs, have been ineffective and politically manipulated, exacerbating water inequality[10].

The P1MC framework prioritizes community mobilization and empowerment. It involves communities in program implementation, fostering ownership, and reinforcing the understanding that access to water is a basic right. This program actively encourages communities to shape and implement initiatives suited to their needs, emphasizing their empowerment and the program’s participatory approach. Social and community participation is integral to the program, beginning with the establishment of municipal committees responsible for oversight and coordination. These committees, comprising various social organizations, provide legitimate channels for community input and ensure accountability in program implementation[8].

According to the Brazilian Government, 1,146,210 cisterns had been constructed by 2022. During periods of increased funding for the P1MC, such as between 2013 and 2015, over 100,000 cisterns were built annually. However, the program has experienced substantial budget cuts since then, with only 5,946 cisterns constructed in 2022, as illustrated in Figure 1.

**Figure 1.**
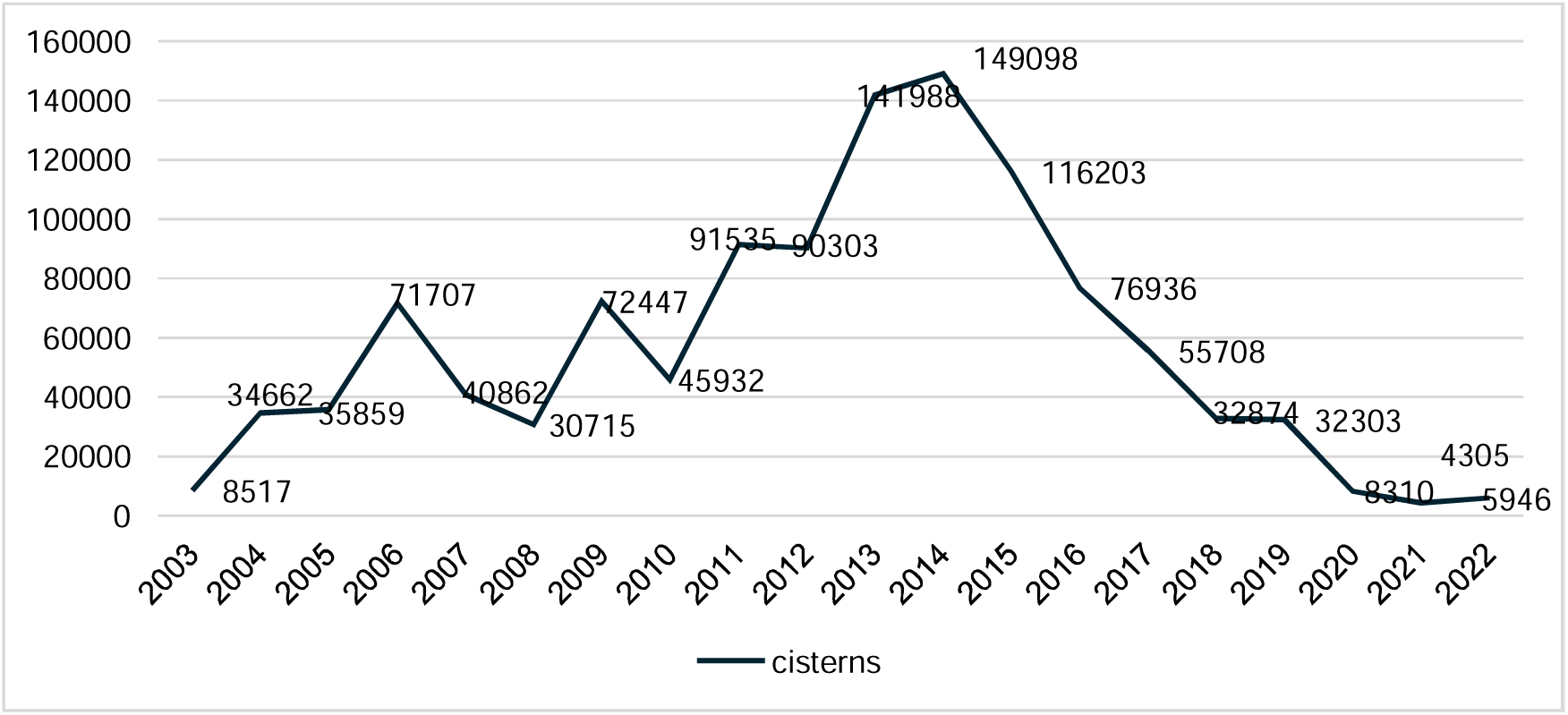
Total number of cisterns built since 2003 in Brazil Source: Brazil, 2023[11].

The P1MC improved water access in Brazil’s Semi-arid region, particularly as climate change exacerbates water scarcity and its associated socioeconomic and health challenges[12,13]. Integrating longitudinal cohort data provides an opportunity to rigorously evaluate the program’s impact on key health outcomes [14], such as child mortality and migration. As climate change drives extreme weather events globally, the P1MC policy becomes even more critical for addressing immediate water access and mitigating broader public health and social vulnerabilities. This evidence-based approach can inform scalable climate adaptation strategies worldwide, particularly in regions facing similar environmental and economic pressures.

Finally, this study evaluates the P1MC climate adaptation policy in Brazil’s semi-arid region and its impact on human health. We synthesize existing literature and, employing a conceptual framework, we identify potential health consequences from climate-induced ecosystem changes and policy-driven adaptation to drought. The DPSEEA model enabled us to trace and understand the context of this specific study on the P1MC public policy and its effects on the health and well-being of the population in the Brazilian Semi-arid region, providing a perspective and a guide for future studies and steps involving this topic. Additionally, we present the cohort of 130 million Brazilians[14] as a significant resource for future research that provides a robust data foundation to explore the associations between social and environmental programs and various health outcomes.

## 2. Methods

### 2.1 Literature search

We searched using web resources such as *Web of Knowledge, Google Scholar, Scielo, Scopus, PubMed,* and *ScienceDirect,* combining terms related to the environmental and health fields. Health-related terms included “health,” “water-related diseases,” “mortality,” and “epidemiology”; environmental terms included “Brazilian Semi-arid region” and “drought,” and our exposure term was the “One Million Cisterns Program.” Additional complimentary terms included “qualitative methods,” “statistical methods,” and “literature review.” We identified approximately 39 studies (scientific articles) describing the P1MC, and only 8 of them associate it with health outcomes. It is important to note that this intervention was specifically designed for the Brazilian Semi-arid region, given its unique characteristics related to natural and climatic issues, as well as the social vulnerability of its population.

### 2.2 The DPSEEA framework

Different approaches and conceptual models from various fields can be used to analyze and manage environmental impacts on human health. Hambling, Weinstein, and Slaney[15] evaluated 11 frameworks for developing environmental health indicators for climate change and indicated that the DPSEEA is the most comprehensive model due to attributes such as conceptual clarity and scope, flexibility, balance between environmental and health issues, and usability. Additionally, the framework’s multidisciplinary approach, conciseness, and ease of communication for decision-making are also cited as advantages.

The DPSEEA (*Driving Force–Pressure–State–Exposure–Effect–Action*) framework, initially developed by the World Health Organization[16,17], has proven its adaptability in various contexts. Morris *et al.*[18] further enhanced this framework to account for the complex impact of contextual factors on the relationship between environment and health. This adaptability is evident in its application in diverse contexts, such as the dynamics of human health and the water environment[19], the implications of changes between natural ecosystems and associations with health services[20], and the association between natural ecosystems, green spaces, climate adaptation and mitigation tools, and health effects[21].

The framework facilitates a structured analysis of the complex interdependencies among environmental changes, societal responses, and health outcomes, highlighting the influences of regional and global environmental drivers[17],[19,20,21]. Additionally, it supports the creation and identification of indicators to aid the monitoring and evaluation of social policies.

The DPSEEA framework is applied systematically to understand the pathway from environmental determinants to health outcomes. First, driving forces, such as socio-economic and demographic factors, are identified using literature reviews or data analysis. Pressures, such as emissions or waste, can then be assessed through quantitative data collection. The state of the environment is evaluated using environmental quality monitoring and spatial analysis. Exposure is measured through modeling or surveys to determine how populations encounter environmental hazards. The effects on health are assessed using epidemiological studies and statistical analysis to establish the relationship between exposures and health outcomes. Finally, policy analysis and stakeholder evaluations identify actions to mitigate these effects. Various applications have been conducted, such as a study on managing healthcare waste[22] and another on drought, leptospirosis, and scorpionism[23].

## 3. Findings

### 3.1 Literature findings

Despite the importance of the P1MC, a significant gap remains in our understanding of its effects on human health. Our literature search identified only eight articles that describe cistern implementation through P1MC, linking it to health outcomes. Luna et al.[24] and Marcynuk et al.[25] found a substantial reduction in diarrheal episodes among households with cisterns compared to those without cisterns. Luna et al.[24] observed a 73% decrease in diarrhea cases, especially among individuals aged 1 to 59, with a more substantial protective effect seen among males (76%). Marcynuk et al.[25] reported a markedly lower 30-day prevalence of acute diarrhea in households with cisterns, mainly among children under five. Both studies noted limitations, including potential selection bias and a lack of mechanisms to improve rainwater quality, suggesting room for improvement in program implementation.

Cisterns have been shown to reduce the prevalence of parasitic infections and diarrheal diseases, as Fonseca et al.[26] found that children from households with cisterns had a lower prevalence of *G. duodenalis* infections (4.8%) than those relying on unsanitary water sources, such as rivers and springs (16.7%). The study emphasizes the importance of water access in reducing parasitic diseases while also highlighting the need for complementary sanitation interventions to mitigate health risks further. In a groundbreaking study, Da Mata et al.[27] demonstrated that access to cisterns during early pregnancy increased birth weight by an average of 1.73 grams per week.

Studies have also pointed out the broader perception of P1MC’s impact on health and welfare. Passador and Passador[13] traced the evolution of public policies in Brazil’s Northeast, showing that cisterns improved the quality of life for families by enhancing health, leisure, income, and gender equality. Similarly, Gomes and Heller[12] found that the program reduced the time spent fetching water by nearly 90%, although challenges related to water quality and socioeconomic conditions persisted. Finally, Fagundes et al.[28] highlighted the positive impact of cisterns on food security and agricultural production. Despite facing high rates of excess weight, cardiovascular risks, and food insecurity, 75% of families involved in the study reported improvements in food security due to the water provided by cisterns. Table 1 summarizes the main aspects of the research discussed.

**Table 1.** Summary of articles describing the impact of cisterns on health outcomes in the Brazilian Semi-arid region.

| Reference<br>and | Data and<br>Methods | Health<br>outcome | Main<br>contribution | Main Results |
| --- | --- | --- | --- | --- |

| location |  |  |  |  |
| --- | --- | --- | --- | --- |
| Luna *et al.*[24]. 21 municipalities of Pernambuco state. | Data was gathered from over 60 days and included 1,765 individuals. Longitudinal prospective study, nested in a cross-section study comparing two groups (households with and without water-tanks). | Occurrence of diarrheal episodes | The findings of this study point to the importance of access to drinking water for the reduction of disease. | Among the 949 individuals with water tanks, there was a reduction in the risk of the occurrence of episodes of diarrhea by 73% compared with the 816 individuals without watertanks (RR=0.27; p<0.001). |
| Marcynuk *et al.*[25]. Agreste Central Region of Pernambuco State | Logistic regression using a face-to-face survey with 3,679 people from 774 households. | Determine the 30 days of prevalence of diarrhoea | This indicates that using cisterns for drinking water is associated with a decreased occurrence of diarrhoea in this study population. Further research | People from households with a cistern had a significantly lower 30-day period prevalence of diarrhoea (prevalence = 11.0%; 95% CI |
|  |  |  | should be conducted to account for additional risk factors and preventative factors. | 9.5-12.4) than people from households without a cistern (prevalence = 18.2%; 95% CI 16.4-20.0). |
| Silva, Heller and Carneiro*[29]. Two municipalities in the Médio Vale do Jequitinhonha, Minas Gerais: Berilo and Chapada do Norte | Quasi-experimental, using survey-based analysis with 664 children under 5 years old. | Diarrhea has occurred within the last 72 hours. | The authors highlight the need to improve the sanitary practices of the rural population so that personal and household hygiene and hygiene concerning consumed water are incorporated into routine habits. | The total prevalence of diarrhea was 5%, but no significant difference between the groups. |
| Fonseca et al.[26]. Berilo and Chapada | Quasi-experimental study cohort with | The prevalence of *G. duodenalis* in | The study suggests the necessity of | They found a higher risk of *G. duodenalis* |
| do Norte<br>(Minas Gerais<br>state) | 664 children. | children under 5<br>years. | complementin<br>g physical<br>interventions with<br>personal and<br>domestic hygiene<br>actions to further<br>reduce<br>parasite<br>infections, which<br>mainly affect<br>underprivileged<br>populations. | infection in<br>children who did<br>not have access to<br>rainwater cisterns<br>compared to those<br>who did (OR 1.72;<br>95% CI 1.14–<br>2.59). |
| Da Mata *et al.*[27]. All<br>Semi-arid<br>region | Administrativ<br>e data from<br>P1MC,<br>CadÚnico, and<br>SINASC. Fixed<br>effects panel<br>regression<br>models. | Birth weight | The paper<br>examines the<br>impact of in-utero<br>exposure to a<br>large-scale<br>climate<br>adaptation<br>program on birth<br>outcomes. | Access to<br>cisterns during<br>early pregnancy<br>increased birth<br>weight,<br>particularly for<br>more educated<br>mothers. We<br>demonstrate that<br>each additional<br>week of in-utero<br>exposure to<br>cisterns is associated with a positive effect on average birth weight, approximately 1.7 grams. |
| Passador and Passador[13]. Municipality of Juazeiro, Bahia. | 34 interviews with benefited families. | Beneficiaries' perception of improved health. | Families reported that water-related illnesses, such as diarrhea, vomiting, and cramps, were quite common in families when they consumed poor-quality water of dubious origin. However, since using water collected in the cistern, such diseases occur very sporadically. | Using cisterns positively influences the quality of life of these families in terms of health, free time, income, and gender issues. |
| Gomes and | Survey-based | Beneficiaries | Emphasize that | Improvements |
| Heller[12]. 63 municipalities of Minas Gerais | analysis with 623 beneficiaries | ' perception of improved health | public water supply policies in Semi-arid rural areas must associate technical issues with management elements that consider local social, climatic, and economic specificities. | in health perception benefited families and reduced the time spent searching for drinking water; however, significant challenges remain in precarious and socioeconomic conditions, as well as those related to an adequate supply of water in terms of quantity and quality. |
| Fagundes *et al.* *[28]. Alagoas state | Survey-based analysis with 29 families. | Food and nutritional security | This study highlights the importance of water access programs for food production within public policies | The Food Insecurity Scale revealed that food insecurity affects 75% of these families. However, focus |
|  |  |  | to guarantee FNS. | groups revealed that families have a positive perception of Boardwalk Cisterns for their food security. They believe that agricultural production has improved, offering a wider range of foods and improving food security. |

Source: Authors own. \*Not focused on the PIMC

### 3.2 The conceptual framework analysis

In this model, climate change is the primary driver exerting various pressures on the ecosystem. These include changes in temperature and precipitation patterns, which are particularly impactful in Brazil’s Semi-arid regions. The pressures in this scenario are multifaceted. Increased frequency and intensity of droughts strain water resources and agricultural productivity; desertification exacerbates land degradation, reducing arable land and affecting biodiversity[23], [30]; increased demand for water due to economic development adds pressures to existing water supplies; economic development drives land use changes, potentially leading to increased pollution and habitat loss[31].

The state of the adaptative ecosystem in the Brazilian Semi-arid region is shaped by its inherent capacity to adapt to environmental stresses. This adaptability is reflected in several specific functions and attributes that are critical for both ecological balance and human well-being. Firstly, the ecosystem supports essential services, such as drinking water and food production, through the cultivation of drought-resistant crops and effective livestock management practices adapted to arid conditions[32,33]. These agricultural practices are vital for local food security. It is interlinked with maintaining nutrient cycling and soil fertility, crucial for sustaining agriculture in nutrient-poor soils prevalent in Semi-arid areas.

The cultural values embedded within these communities, manifested in traditional knowledge systems related to water management and sustainable living, are components of the ecosystem state. These practices include ancient techniques for water conservation, such as the construction and use of dams, weirs, and cisterns, as well as soil preservation methods that have been passed down through generations. These cultural practices are not only survival strategies but also key to maintaining ecological balance and enhancing the community’s and ecosystem’s resilience to climatic adversities [34,35,36].

Regarding the contextual factors, the convergence of limited infrastructure, socioeconomic disparities, cultural practices, and acute water shortages creates a multifaceted exposure that critically impacts community health and resilience. Infrastructure deficiencies – ranging from inadequate water supply systems to limited access to healthcare – intersect with economic inequalities, further stratifying communities based on their ability to adapt to environmental pressures. Lower-income groups often lack the resources to implement effective water conservation techniques or access emergent agricultural technologies essential for survival in arid conditions[37,38].

Culturally, the ingrained traditional knowledge of water management and land use shapes community responses to these environmental challenges[38]. However, the effectiveness of combining traditional practices with modern adaptation strategies varies significantly across different communities, influenced by cultural acceptance and the practicality of integrating new solutions into established lifestyles. For instance, traditional rainwater harvesting methods might be enhanced by modern cisterns, yet acceptance varies based on perceived benefits and the alignment with cultural values regarding land and water stewardship.

Furthermore, the challenge of water scarcity serves as a catalyst that exacerbates existing infrastructural and socioeconomic vulnerabilities. It influences public health outcomes through increased incidences of waterborne diseases and conditions associated with poor nutrition and sanitation. The resultant decline in crop yields exacerbates food scarcity, leading to an upsurge in malnutrition-related illnesses. This situation is particularly dire for vulnerable populations, such as children and the elderly, who face increased risks of anemia, vitamin deficiencies, and other diet-related health issues[28].

As droughts become more severe, water scarcity leads to an increased reliance on potentially contaminated water sources. This rise in water contamination directly contributes to an increase in waterborne diseases, such as diarrhea, cholera, and various parasitic infections, as populations resort to less reliable water sources for daily use [28,29,32].

The psychological impact of these environmental stresses also becomes profound. The persistent threats of water scarcity, food insecurity, and economic instability cultivate a pervasive sense of uncertainty and stress among communities. This psychological burden can lead to increased incidences of anxiety, depression, and other mental health disorders, compounding the community’s health challenges[39].

**Figure 4.**
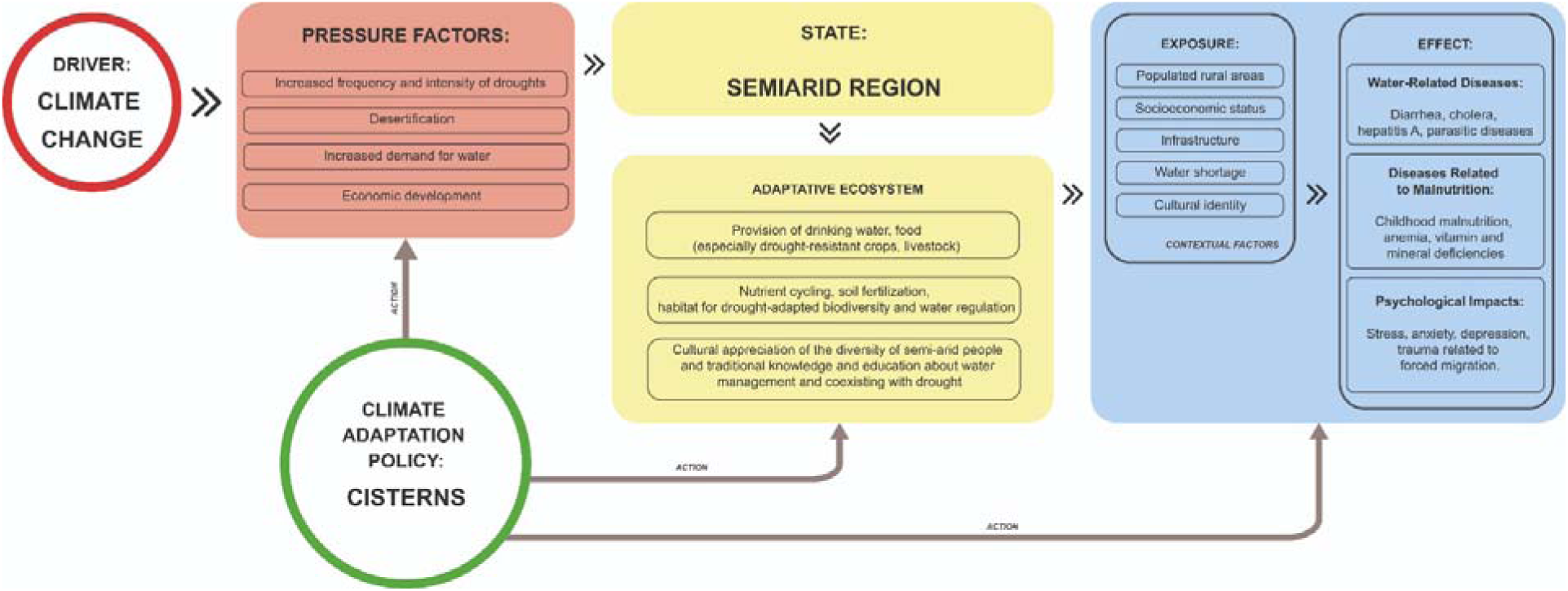
The conceptual analysis of the P1MC based on the DPSEEA framework Source: Adapted from WHO (1999).

Finally, implementing cisterns as a social technology counters the pressures of increased drought frequency and desertification in a scenario of intensification of climate extreme events by providing a stable and sustainable water supply. This technology directly mitigates water scarcity by capturing and storing rainwater during rainy seasons for use during droughts. Water availability supports the adaptive ecosystem, sustaining human consumption during dry periods. The strategic placement of cisterns influences various exposure variables and contextual factors that affect the local populations. When enhancing water security, cisterns alleviate socioeconomic disparities and equip even the most economically disadvantaged communities with the means to collect and store water. This addresses infrastructure gaps and aligns with traditional water management practices, ensuring cultural integration and acceptance.

## 4. Discussion

The studies on the P1MC demonstrate significant health improvements in Brazil’s Semi-arid region, specifically a reduction in diarrheal episodes in households with cisterns, improved birth weights, and enhanced food security of participating families reporting positive impacts on agricultural productivity[24,25,27,28]. These outcomes are linked to increased access to clean water, underscoring the importance of water security as a determinant of health. Although there is still little evidence, these findings have global relevance, particularly for regions facing similar climate-induced water scarcity challenges, such as Sub-Saharan Africa and South Asia. Implementing cisterns mitigates the health risks associated with waterborne diseases, undernutrition, and poor maternal and child health by providing a stable, local water supply.

Applying P1MC as a climate adaptation technology demonstrates its potential for scalability and relevance in global health interventions for drought-prone regions. The need for studies evaluating the health impacts of the P1MC represents a significant gap in literature, given the growing relevance of water security in the face of climate change. It highlights the urgent need for a comprehensive research agenda focused on evaluating the health outcomes of P1MC. Health impacts are profoundly affected by the use of cisterns[27]. Reliable access to clean water can decrease the incidence of waterborne diseases, which are prevalent during periods of water scarcity when populations may otherwise rely on contaminated sources[25]. Furthermore, irrigating crops with stored rainwater can mitigate the effects of malnutrition by increasing food availability, even during prolonged droughts [40]. This dual benefit of cisterns – improving water and food security – enhances communities’ overall health and resilience.

The application of the DPSEEA framework also facilitates the identification of indicators to support assessments of climate change and adaptive strategies in the Brazilian Semi-arid region. This exercise, which does not aim to exhaust the number of indicators, will differ from other applications by adding a qualitative dimension, broadening the scope of information, and incorporating the experiences, perceptions, and perspectives of people who experience the limitations and adaptations in a Semi-arid environment affected by climate change. We understand that incorporating social participation into the knowledge production process is an important strategy for better connecting with problems and solutions and expanding the potential applicability of the results. Klenk *et al*.[41] demonstrate how climate change and health research have incorporated local knowledge, albeit with limitations, such as a focus on people’s perceptions.

In our framework, we identify quantitative indicators, including rates of temperature increase, precipitation, duration and frequency of droughts, water reserves, sociodemographic data, mortality rates, and disease prevalence. In addition, we have listed some qualitative indicators that will help capture the complexity of human experience in this environment. These include changes in hygiene practices, perceptions of water quality, and reports of psychological stress. When activating these subjective components, different methodologies will be required in the analyses, such as focus groups, interviews, and participant observation. Table 2 lists the indicators by framework level and provides a preliminary exercise of recommended actions that should be validated and complemented with the collaboration of participants living in the Semi-arid region in the next stages of this study.

**Table 2.** Qualitative indicators supporting action on P1MC.

| Level | Determinants or drivers | Indicators | Actions |
| --- | --- | --- | --- |
| Driver | Climate change | Increase rate of temperature | Development and monitoring of a heat-health action plan for the Semi-arid region. |
| Pressure factors | Increased Frequency and intensity of droughts | 1. Precipitation level<br>2. Drought duration and frequency | Monitoring to prevent deforestation. |
|  |  | 3. Water reservoir levels | Environmental education on how to avoid soil pollution.<br>Support for family farming.<br>Social cash transfer program. |
|  | Desertification | 1. Decline vegetation area between time intervals<br>2. Decrease in soil organic matter content between time intervals |  |
|  | Increased demand for water | 1. Number of water trucks |  |
|  | Economic development | 1. GDP<br>2. Poverty rate<br>3. HDI |  |
| State | Semi-arid region | 1. Region |  |
| Exposure | Populated rural areas | 1. Population in drought areas<br>2. Rate of migration<br>3. Number of children, adults and elderly (cohort data) | Educational Policy for Youth in the Territories.<br>Sustainable economic development policies.<br>P1MC improvements.<br>Health promotion actions and educational materials to preserve technology |
|  | Socioeconomic status | 1. Income rates<br>2. Education rates<br>3. Access to healthcare<br>4. Age<br>5. Gender<br>6. Race-color (cohort data) |  |
|  | Infrastructure | 1. Water coverage and sources |  |
|  | Water shortage | 1. Number of houses without cisterns<br>2. Time to access water |  |
|  | Cultural identity | 1. Changes in hygiene practices<br>2. Building capacity in managing cistern water |  |
|  | Water quality | 1. Perception of water quality by beneficiaries<br>2. Water indicators (pH level, turbidity, etc.)<br>3. Frequency of water quality tests |  |
| Effects | Water-related diseases: Diarrhea, cholera, hepatitis A, parasitic diseases | 1. Disease rates<br>2. Mortality<br>3. Hospitalization | Health surveillance and promotion programs<br>Improvements of PHC |
|  | Diseases related to malnutrition: Childhood malnutrition, anemia, vitamin and mineral deficiencies | 1. Malnutrition diseases rates<br>2. Morbidity and mortality caused by these diseases<br>3. Food diversity index (cohort data) |  |
|  | Psychological impacts: stress, | 1. Disease rates<br>2. Behavior changes |  |
|  | anxiety, depression,<br>and trauma related<br>to forced migration | 3. Testimonies on<br>psychological distress<br>(cohort data) |  |
|  | Health and Well-Being | 1. Perception of life<br>satisfaction<br>2. HDI |  |

Source: Authors' own.

In Semi-arid regions, inconsistent rainfall patterns mean families may struggle to maintain and refill cisterns during extended dry spells, potentially limiting the reservoirs’ ability to provide clean water consistently. This is further complicated by the economic constraints many households face, as maintaining and managing these cisterns often require additional financial and logistical resources that low-income families may not readily have access to. The costs associated with water treatment, maintenance, and sometimes even purchasing water during droughts can undermine the long-term sustainability of such programs.

Similar challenges have been addressed in other settings through innovative approaches that help families maintain water security despite variable rainfall[42]. For example, in Sub-Saharan Africa and South Asia, rainwater harvesting systems are often supplemented by community-managed water storage facilities or water delivery programs during dry periods[43,44]. In Ethiopia’s Tigray region, water-sharing cooperatives enable multiple households to pool their resources for the upkeep and maintenance of water storage facilities, thereby reducing individual economic burdens[45]. Financial support mechanisms, such as subsidies or microloans for water infrastructure maintenance, have been implemented in parts of India to enable low-income families to manage their cisterns effectively[46].

Similar solutions could be adopted to overcome the limitations of rainfall variability in the P1MC. Introducing community-managed water reserves or establishing contingency funds to help families maintain and refill cisterns during droughts would increase the program’s resilience. Furthermore, government policies that subsidize water treatment supplies or offer financial aid to low-income families could alleviate the economic strain and ensure sustained access to water, even in times of scarcity. By learning from these global examples, the P1MC can be strengthened to withstand the challenges posed by climate-induced water scarcity.

### 4.1 The role of cohort data in the assessment of climate change impact and the development of climate adaptation strategies

To assess the impact of climate and climate adaptation strategies on health, we propose utilizing the Center for Data and Knowledge Integration for Health (CIDACS) 100 Million Brazilian Cohort as a sizable, secure database with immense potential for studies on climate change, adaptation policies, and their various effects on human health. The 100 Million Brazilian Cohort represents a groundbreaking initiative in public health research in Brazil, developed in response to the scattered yet high-quality social and health databases available in the country[14]. This cohort was established to integrate these databases and assess the impact of various social protection policies on health outcomes. The overarching goal is to improve understanding of how these policies influence the social determinants of health overall and within specific subgroups of interest over time. From 2001 to 2018, the cohort’s baseline amassed data on 131.7 million low-income individuals from over 35 million families[14]. This population is predominantly comprised of children and young adults, with a larger proportion of females than the general Brazilian population. The 100 Million Brazilian Cohort provides a unique opportunity to advance the study of climate adaptation policies in the Brazilian Semi-arid region. This vast cohort presents several key advantages that can be leveraged to understand and enhance the effectiveness of interventions aimed at mitigating the impacts of climate change in this vulnerable area. We can enumerate some key points such as:

1. *Comprehensive and longitudinal data:* The Cohort’s integration of health and social data from various government sectors enables the exploration of the social determinants of health and their interplay with environmental factors. The longitudinal nature of the data is especially valuable, with follow-ups beginning in 2001 and continuing through the period following the implementation of numerous social policies in 2003. It enables analysis of the lasting effects of climate adaptation strategies, such as the introduction of water-saving technologies like cisterns, on health outcomes over time.
2. *Sub-population analyses:* Given the Cohort’s large size, researchers can study less common health outcomes and their variations across different sub-populations, including those most affected by climate change, such as rural, indigenous, and isolated communities. This capability is critical in the Semi-arid region, where diverse ethnic and socioeconomic groups may experience the effects of climate change differently.
3. *Detailed interaction studies:* The Cohort enables the examination of detailed interactions among various factors, such as age, gender, and race, as well as the effects of combined social and climate-related policies. It can help to elucidate how different demographic groups respond to climate adaptation measures and which strategies are most effective in improving health outcomes in the context of ongoing environmental changes.
4. *High data quality and reduced bias:* Using administrative data minimizes the recall bias often associated with self-reported data, thereby enhancing the reliability of studies on service usage and health outcomes. The linkage techniques and continuous quality checks ensure the accuracy of data, which is important for forming robust inferences about the effectiveness of climate adaptation interventions.
5. *Focus on vulnerable populations:* Since the Cohort includes a significant representation of the poorer segments of the population, it is well-suited to explore the impacts of climate adaptation policies on those most vulnerable to climate change. This focus is particularly relevant in the Semi-arid region, where economic challenges and limited access to resources compound the effects of environmental stressors.

The 100 Million Brazilian Cohort provides data that allows for the analysis of climate-related health risks in the Semi-arid region of Brazil. This evidence supports the development of adaptation policies that consider regional characteristics. It can inform decision-making at local and national levels to improve health outcomes in populations highly exposed to climate change.

## 5. Conclusions

As a core component of the P1MC, the cisterns stand out as a culturally resonant adaptation tool that embodies the intersection of environmental necessity and local cultural practices. As evidence suggests, cisterns have influenced the health and well-being of local populations, proving crucial in drought by providing necessary water reserves. To this end, it is important to highlight that ASA’s engagement has the potential to support the implementation of the social technology of cisterns in other territorial contexts.

However, while cisterns are vital resources, their effectiveness is limited by several factors, including rainfall variability and families’ economic capabilities to maintain and refill these reservoirs during prolonged dry spells[27]. Maintenance issues further complicate their functionality, affecting their long-term viability as a sustainable water source. These limitations emphasize the need for comprehensive policies that not only support the installation of cisterns but also ensure their maintenance and the continuous training of communities to manage their water resources efficiently.

Harnessing the comprehensive and detailed data from the 100 million Brazilian Cohort will allow us to understand the dynamics within Brazil’s Semi-arid region and foster the development of evidence-based climate adaptation strategies. Integrating this extensive dataset with records from P1MC and *CadÚnico* provides insights into the effectiveness of interventions, such as cistern installation, which directly correlate with improved health outcomes and a better quality of life for vulnerable populations. Cisterns have proven to be effective during periods of severe drought, highlighting the direct impact of targeted adaptation policies. Furthermore, the ability to track health outcomes such as child mortality and diseases related to water quality provides a clear indicator of success and areas for improvement in current policies. To continue leveraging this cohort data, policy decisions can be better informed at all levels, ensuring that adaptation strategies will address the immediate needs of these communities and contribute to sustainable development and resilience in one of the regions most susceptible to the adverse effects of climate change.

As we move forward, policymakers must consider the successes and limitations of current strategies, adapting and evolving policies better to meet the needs of Brazil’s semi-arid populations. A holistic approach will ensure that the benefits of programs like the P1MC are maximized, contributing to the sustainability and well-being of vulnerable communities in the face of ongoing climate challenges. Finally, when producing a contextualized study, it is crucial to emphasize that achieving listening and understanding life beyond data, engaging the community and policymakers, is essential to enhance a public policy like P1MC and improve the health of communities that need support to thrive in their specific contexts.

## Supporting information

https://github.com/FirminoFilho/Supplement-P1MC-case-study/blob/main/Supplement.pdf

## Acknowledgment

This manuscript is part of a supplement titled Lessons from the Field: Case Studies *to advance research on climate adaptation strategies and their impact on public health.* This writing project was supported by the National Institutes of Health (NIH) Climate Change and Health Initiative (https://climateandhealth.nih.gov) and coordinated by the Center for Global Health Studies at the Fogarty International Center of NIH. A steering committee of global experts on health and climate change led the activity. More information is available at https://go.nih.gov/ClimateAdaptationStudies.

This research was also funded by the NIHR (NIHR134801), using UK aid from the UK Government to support global health research. The views expressed in this publication are those of the author(s) and not necessarily those of the NIHR or the UK government.

## Data statement

All data used in this study are public available.

