## Supplementary material for "Water, Health, and Social Technologies: One Million Cisterns Programme case study": https://github.com/FirminoFilho/Supplement-P1MC-case-study/blob/main/Supplement.pdf

**SM1. Climate change, Semi-arid and water scarcity: a territorial perspective of the P1MC**

Historically, drought in the Semi-arid region has been intertwined with social and economic hardships, often exacerbating the vulnerability of poor populations. One of the first well-documented droughts in Brazil was the great drought of 1877-1879, a global event that affected territories such as Brazil, India, Korea, the Philippines, Java, the Maghreb, New Caledonia, Northeast China, Southern Africa, and the Mediterranean. According to Secreto[1], the absence of support led to three global-scale droughts (1877-1879, 1889-1891, and 1896-1902), resulting in profound crises that hindered people's survival and caused an estimated loss of approximately 30 million lives.

The climate in the region is characterized by irregular rainfall and high evapotranspiration rates, which contribute to a constant risk of water scarcity. Both the absence or scarcity of rainfall, as well as its high spatial and temporal variability, contribute to the occurrence of droughts. Semi-arid - a natural and cyclical phenomenon in the region. The soil is predominantly crystalline, which limits the storage of sufficient water in underground sources (aquifers). The shallow depth of the soil also reduces its capacity to absorb rainwater, contributing to droughts. Furthermore, approximately 90% of rainwater cannot be used due to the high evaporation rate in the Semi-arid region and its surface runoff due to the crystalline soil. Average temperatures are high (above 26ºC), and it is natural in Semi-arid areas for the volume of rain to be lower than the evaporation rate[2].

For a municipality to be considered part of the Brazilian Semi-arid region, one of the following technical and scientific criteria must be met [2,3]: i. average annual rainfall equal to or less than 800 mm; ii. Considering the last few decades, the Thornthwaite Aridity Index should be equal to or less than 0.50; iii. daily percentage of water deficit equal to or greater than 60 % considering all days of the year. The aridity index indicates a region's water deficiency based on precipitation, evapotranspiration, and the available water capacity in the soil [4]. The Brazilian delimitation of the Semi-arid region is not fixed. The climate behaviour of the last decade, marked predominantly by severe droughts, reinforces the change in the outline of the Brazilian Semi-arid region, and more municipalities have been added to the Semi-arid climate over the years.

The drought phenomenon cannot be considered in isolation from its social context, as the water crisis generated a governmental crisis characterized by land concentration and delays in decision-making, as well as the creation of public policies to mitigate problems that extended beyond the drought[1]. In recent years, the impacts of climate change and desertification have intensified these challenges, further threatening the livelihoods of those in the Brazilian Semi-arid region[5,6,7].

The Brazilian government had implemented various strategies to adapt to life's challenges in the Brazilian Semi-arid Region. However, the continuous waves of emigration from this area throughout the 20th century highlight the ineffectiveness of these efforts, which included the construction of large-scale water reservoirs and the establishment of numerous support agencies. Even though these measures have enhanced the region's water storage capacity, they have not adequately addressed the issue of water access for the impoverished rural population[8].

Before implementing public policies related to rainwater harvesting in the Semi-arid region, populations living in rural or sparsely populated urban areas had to travel considerable distances to reservoirs, commonly referred to as *açudes* (weirs), to find water for drinking and hygiene needs. This task was predominantly undertaken by women and children[9,10,11]. It is important to note that the concentration of water in a few reservoirs, many of them private despite being built with public funds, exacerbated the challenge of accessing water[12].

In the 1990s, despite limited resources from non-governmental organizations or personal family funds, the construction of cisterns gained momentum in the interior of the Northeast. Families with purchasing power from agriculture, cattle farming, or sheep farming could trade their produce and allocate resources to build cisterns adjacent to their residences. Once completed, these cisterns served as water reserves for a limited period, as the stored water was often shared with neighboring families who lacked the resources to construct their cisterns. Cooperation among families in the Semi-arid region was more than a one-time aid; it was a survival strategy. Thus, when the water ran out, families would return to fetch water from nearby reservoirs.

This situation changed significantly with the expansion of the P1MC project, particularly from 2003 onwards[9,13,14]. This public policy was an initiative promoted by social movements and non-governmental organizations before 2003 and received formal support from the federal government starting in 2003. The P1MC receives funding from the federal government, while the ASA contributes expertise in family mapping, cistern construction techniques, and training on water storage and usage. Municipal social assistance agencies collaborate on family mapping. The prioritization of families eligible for benefits is based on both income criteria and proximity to water reservoirs. Currently, the Cisterns Program is regulated by Law No. 12,873, 2013, Decree Nº 9,606, 2018, and a series of ordinances and normative instructions accessible on official government portals.

The initiative has been implemented in nine semi-arid states of the Northeast region of Brazil, covering 86.48% of its area, as well as in parts of the southeastern states of Minas Gerais (11.01%) and Espírito Santo (2.5%)[15]. The primary aim of the policy is to drive social transformation through education, empowering vulnerable populations by providing water and fostering new social relationships that break away from the traditional patron-client dynamics prevalent in the region. It frames water preservation, access, and management as both a right and a responsibility for all citizens.

In this context, the P1MC promotes a sustainable approach to addressing the challenges of the Semi-arid ecosystem by prioritizing long-term educational measures over purely technical solutions, making it distinct from previous efforts. Local NGOs identified the cylindrical cistern as the most suitable type for the region's soils, being easy to construct, durable, and cost-effective[10,15]. Figures 1a and 1b depict a cistern of P1MC in the Brazilian Semi-arid region constructed in the rural area to store rainwater. Traditionally, these cisterns are made from concrete slabs, and the residences are equipped with suitable water catchment infrastructure to channel water from the roofs through gutters into the cisterns. Beneficiary families actively participate in the construction process, which enhances their sense of ownership and fulfillment and preserves the territorial identity throughout cooperation.

Figure 1. Cistern suitable for water storage

Figure 1a

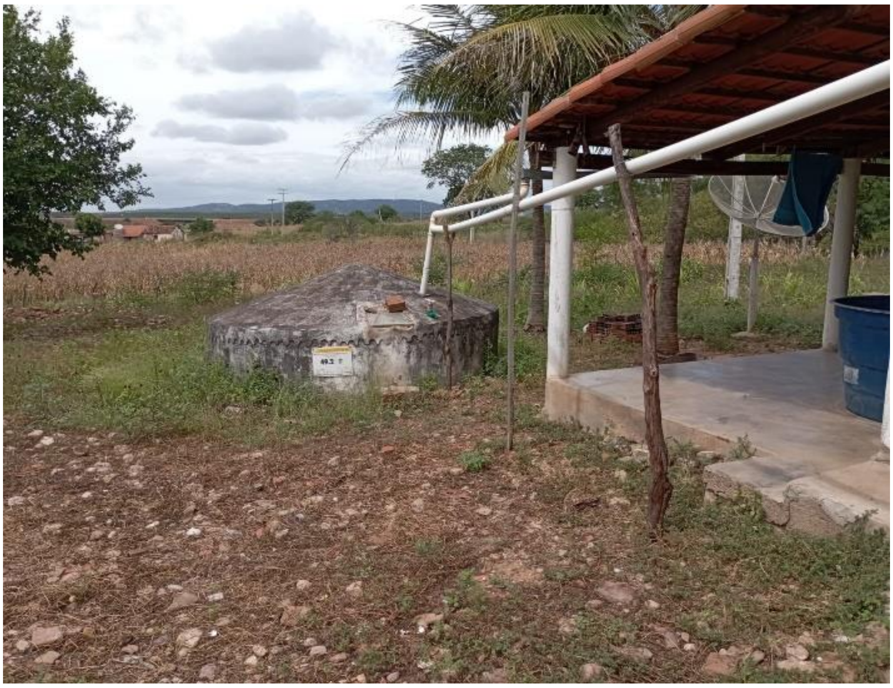

Figure 1b

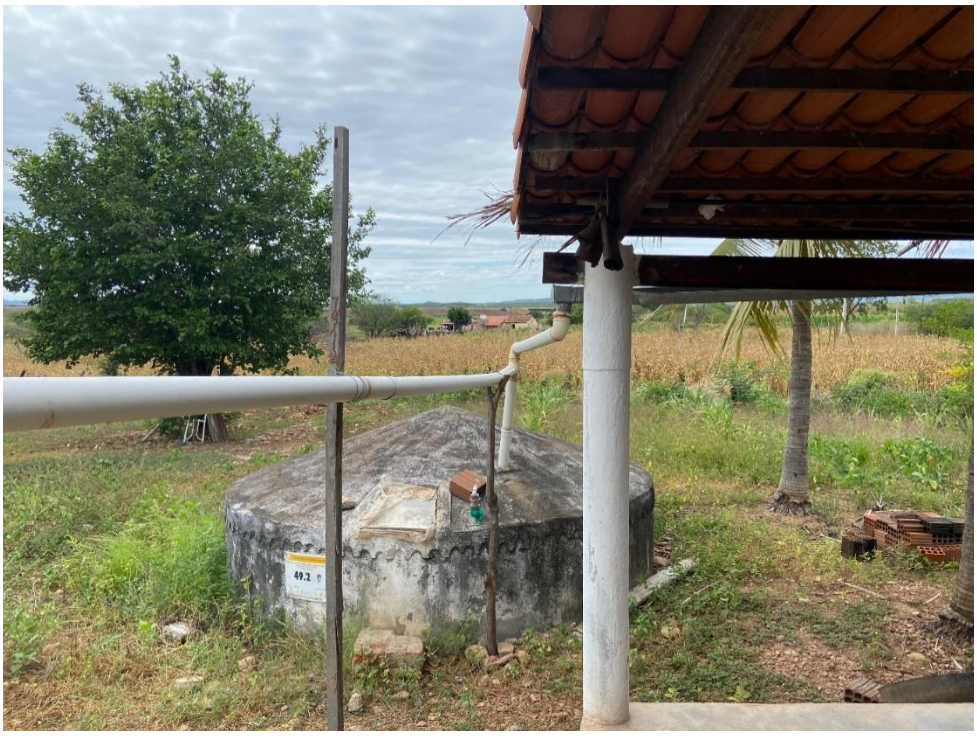

Source: Authors' own.

Furthermore, the implementation of infrastructure in the Semi-arid region faces other challenges due to social inequalities and different cultural contexts. When rainfall is insufficient, additional demands arise, such as the transportation of water by tank trucks from reservoirs suitable for human consumption, as shown in Figure 2. When this demand is not funded by public funding, it requires additional costs from families or communities.

Figure 2. Weir for human consumption

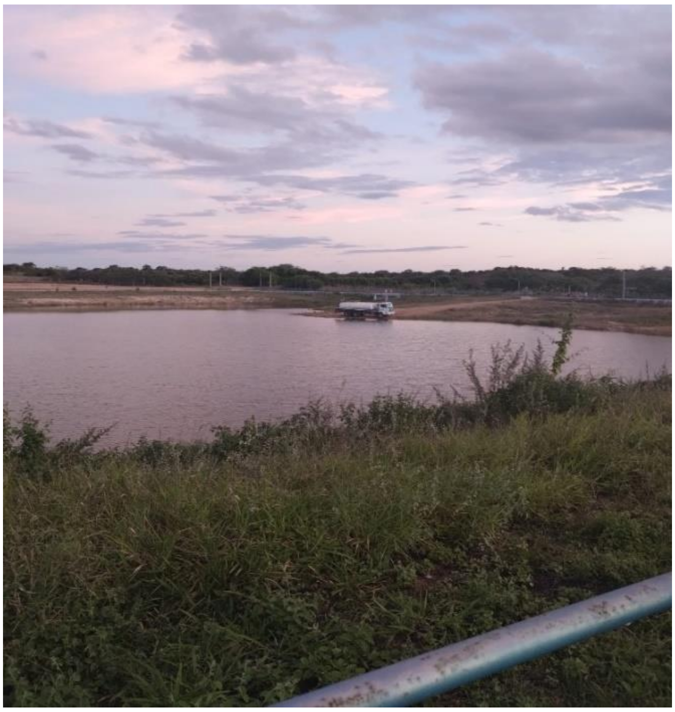

Source: Authors' own.

Additionally, the concentration of water resources in private reservoirs and wells, although water is a basic right of the population, often requires financial contributions from families to ensure continuous access to this essential resource. Regular maintenance of cisterns is crucial to prevent degradation and ensure water quality, which in turn directly impacts the health of the families benefiting from it. The cistern in Figure 3 appears to be in a state of deterioration, surrounded by vegetation, lacking paint, and without the necessary water catchment structure on the residence.

Figure 3. Cistern not suitable for water storage

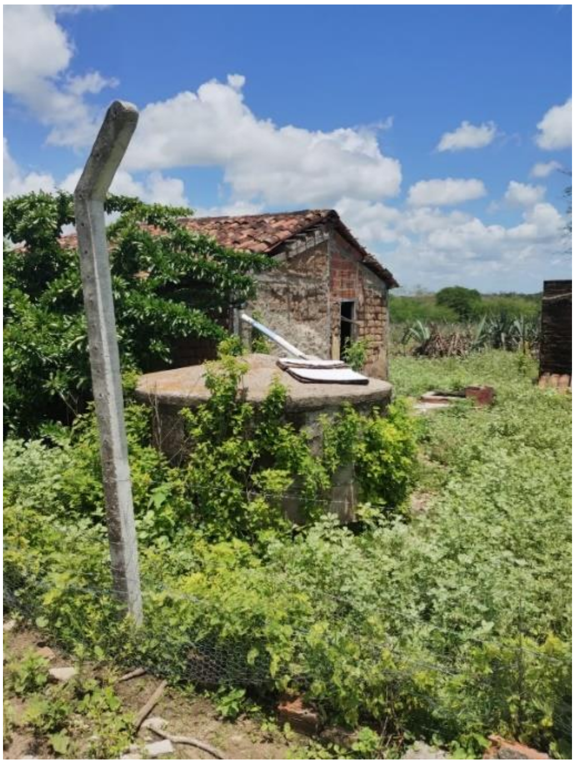

Source: Authors' own.

The educational process is expected to enable the community to participate in and influence public policies for sustainable regional development. A cistern with a 16,000-liter capacity can supply a family of five with water for 10-12 months, reducing the need to travel long distances to fetch water and decreasing their dependency on the local elite, who have historically controlled water resources[16]. The project also provides training in cistern construction for both men and women, offering new income opportunities and enhancing the community's self-sufficiency and resilience[17].

The cisterns program, firmly grounded in the concept of living harmoniously with the Semi-arid environment, serves as a gateway to the creation of a collective social identity. This identity allows individuals to find holistic meaning in their experiences, guiding emancipatory and autonomous processes. The approach is dedicated to expanding the creative capacities of the local population by optimizing existing resources and, importantly, reviving traditional knowledge and practices. This cultural revival supports populations historically vulnerable to drought impacts and also represents a new strategic direction for public intervention, promoting social innovation and sustainable development through social technologies based on the paradigm and social movement known as "coexisting with the Semi-arid" (CSA)[18].

As collective action strengthens, farming families become key protagonists in the process, supported by their own experiences and local knowledge. The emergence of new forms of political governance in the Semi-arid region is a direct result of these social experiences and expectations[18]. It demands a co-responsibility between the state and civil society, as well as between the government and social movements, establishing a novel dynamic centered on common objectives and rejecting dominant forms of political and social organization.

The relationship between the state and civil society around the P1MC, which generated broad expectations, forms the foundation of the paradigm of living harmoniously with the Semi-arid environment. The cisterns program promotes democratizing access to potable water, ensuring food and nutritional security, and encouraging social participation through innovative practices. It establishes mechanisms to overcome clientelism and subordination associated with the “drought industry” [12,19]. From a decolonial ecology perspective, according to Ferdinand[20] it is necessary to consider the fact that the environmental crisis is directly related to Colonialism and its severe wounds like slavery, giving place to environmental racism.

A territorial perspective highlights the intricate connections between water scarcity, climate change, and health in the Brazilian Semi-arid region. When we address the territorial inequalities and promote integrated solutions, we can build a more resilient future for all Semi-arid populations. Climate change is severely impacting livelihoods, particularly in regions heavily dependent on agriculture, leading to economic instability [21]. This economic strain limits families' ability to afford healthcare, nutritious food, and other essentials, thereby worsening health outcomes for children and the elderly [22]. From a gender perspective, women, who often bear the responsibility of caregiving and household management, are facing increased burdens that perpetuate a cycle of poverty and poor health, with significant long-term effects on community well-being.

Communities with strong social cohesion are better equipped to withstand and recover from the impacts of climate change. The presence of robust social support systems is particularly crucial for the health of vulnerable groups, such as the elderly and children [22]. In fragmented communities, the absence of such support can lead to neglect and poor health outcomes. Effective community resilience not only mitigates immediate health risks but also fosters an environment where long-term adaptive strategies can be developed and implemented. To effectively address the health impacts of climate change in the Brazilian Semi-arid region, it is essential to have a comprehensive understanding of how territoriality, socioeconomic factors, and social and cultural dynamics intersect. Empowering vulnerable populations through targeted social technologies and inclusive policies will be crucial for building a more resilient and healthy future in the face of ongoing environmental challenges.
